# A Method to Identify the Missing COVID-19 Cases in the U.S. and Results for mid-April 2020

**DOI:** 10.1101/2020.04.28.20083782

**Authors:** Theodore R. Breton

## Abstract

I use the COVID-19 death rate in South Korea and a method relating the ratio of death rates in a U.S. state to its share of cumulative positive tests to estimate the total cases of COVID-19 in the U.S. and to estimate the extent of infection and the unidentified share of the infected population in each of the lower-48 states and in New York City in mid-April, 2020. I identify a logarithmic relationship between the cumulative death rate in a state and its cumulative positive share of tests. Using this relationship, I find that 4.3-5.4 million people, 1.4-1.7% of the U.S. population, were infected, with rates of infection that ranged from 0.1% in more rural states to 8-10% in New York state and 11-13% in New York City. Only 16-20% of these infected individuals were identified later through testing.

---

Extremely limited testing for COVID-19 until recently and the uncertain share of asymptomatic cases has led to considerable speculation about the extent of infection in the U.S. population. In this paper I use state level data on positive COVID-19 test rates and associated deaths, combined with an estimate of the death rate from the virus in South Korea, to estimate the number of coronavirus cases in each state, in New York City, and in the U.S. in total. The methodology presented in this paper is similar to the methodology in Breton [2020], but the data used in this paper to estimate the share of infection covers a longer period and the interpretation of the model’s results has been revised based on new information.

## I. Methodology

I have developed a methodology to estimate the unidentified cases of COVID-19 in the U.S. that utilizes death rates in South Korea and in the lower-48 states of the U.S. and the share of cumulative positive tests in the lower-48 states. The methodology is based on a series of assumptions:

1. Unlike all other countries, South Korea’s documented experience with the coronavirus is accurate and complete.
2. South Korea and the U.S. are sufficiently similar in their medical treatment capacity to have similar actual death rates (deaths/infected individuals) for COVID-19 for similar age distributions of patients.
3. The reported share of positive tests in a U.S. state or region is an indicator of the share of infected individuals tested, with a higher positive share inversely related to the share of total cases identified by tests
4. The expected death rate in a state associated with its share of positive tests divided by the death rate in South Korea provides a valid ratio of the total cases/identified cases in the state.

I do not use the death rate in each state directly to estimate unidentified cases in the U.S, because state death rates vary based on the particular populations infected in the state (e.g., the elderly in nursing homes). Instead I use the death rates in all of the states to estimate a relationship between the expected death rate and the share of cumulative positive tests in a state. I do this because the share of positive tests in a state is likely to relate more closely to the share of COVID-19 cases that have been identified than the state’s particular death rate.

The critical element in implementing my method is to account properly for the lags between contagion, the onset of symptoms, the reporting of test results, and deaths. The typical lag between contagion and death is 23-27 days [Boyd, 2020]. Due to delays in U.S. testing in April 2020, this means that positive tests for COVID-19 identified infection that occurred 8-12 days earlier and relate to deaths that occurred 11-19 days later.

I use the estimated average lag between the reporting of COVID-19 positive tests and death to identify the related deaths and calculate the death rates in South Korea and in the lower-48 states that correspond to the reported cases in early April. I use an assumed lag of 19 days from the onset of symptoms to death to calculate the death rates. This lag is consistent with the estimate of 18-21 days in the UK [Boyd, 2020] and Verity et al.’s [2020] estimate of 17.8 days in China. I use reports on testing delays to estimate the lag between test results and deaths.

I then plot the death rate in each state vs. the share of positive tests on April 7^th^ and use a logarithmic trendline to determine the relationship between these two variables in the state data. I calculate the ratio between the estimated death rate in the U.S. and in South Korea to determine the relationship between the actual total cases and the reported cases in each state or in a subregion of a state. I use this relationship to estimate the identified and unidentified shares of the total cases in the states on April 25th, which in total provides a national estimate of the infected population in the U.S. in mid-April.

I validate the methodology by examining whether the estimate of the unidentified share of cases in a state is consistent with its reported testing strategy and the availability of testing in March and early April. Since the states did not test asymptomatic individuals, I also examine whether the estimated shares of unidentified cases are consistent with an estimate of the asymptomatic share of all cases in a screening study carried out in Iceland.

In this study 100 of 13,080 (0.6%) participants tested positive and were quarantined for 14 days, during which 43% reported that they were asymptomatic [Gudbjartsson, et al., 2020]. The study excluded individuals with severe symptoms that would be hospitalized. A WHO-China joint mission determined that about 20% of all symptomatic cases in China required hospitalization [Verity, 2020]. Assuming this ratio is applicable in Iceland, I reduced the estimated asymptomatic share to 38% and use this share to further test whether my estimates of the range of unidentified shares of COVID-19 cases in the U.S. are plausible.

## II. The Death Rate in South Korea

The death rate used in this study is the share of positive cases of COVID-19 that end in death. I use data from South Korea to estimate the actual death rate in the U.S.

The government of South Korea implemented a contact tracing strategy to identify all of the individuals infected with COVID-19 in the country, including those with no symptoms, and then tracked these individuals to determine the associated deaths. Table 1 presents the data for the cumulative cases of the virus and the associated deaths in South Korea during April. Since the number of new cases declined to almost zero during the month, there must not have been many asymptomatic individuals who were not tested, identified, and quarantined. This means that South Korea is unique among countries in including all of its asymptomatic cases in the national estimate of total COVID-19 cases.

**Table 1.** South Korea Coronavirus Statistics.

| **Table 1**<br>**South Korea Coronavirus Statistics** |  |  |  |  |  |  |  |
| --- | --- | --- | --- | --- | --- | --- | --- |
|  | Tests Completed | Total Cases | Total Deaths | Population | Positive Test Rate | Death Rate | Infected Rate |
| April 4th |  | 10,156 |  |  |  |  |  |
| April 18th | 541,284 | 10,653 | 232 | 51,600,000 | 2.0% |  | 0.02% |
| April 20th | 551,054<sup>1</sup> | 10,674 | 236 | 51,600,000 | 2.0% | 2.3% | 0.02% |
| Source: Roser, Ritchie, Ortiz-Ospina and Hasell (2020) |  |  |  |  |  |  |  |
| <sup>1</sup>11,981 tests outstanding (2-3 days) |  |  |  |  |  |  |  |

The Government of the Republic of Korea [2020] reports that in early April the delay between testing and reporting results was 2-3 days. I assume 19 days between the onset of symptoms and death and three days between the onset of symptoms and test confirmation, which leaves 16 days as the average lag between confirmed test results and death.

As shown in Table 1, there were 10,156 cumulative confirmed cases of the virus in South Korea on April 4^th^, and there were 236 cumulative deaths from the virus sixteen days later on April 20^th^. This yields a 2.3% death rate for all cases.

This rate could be low if some deaths related to the cases are yet to occur or are not included in the statistics, but this is unlikely. The data in the table show that in the two days prior to April 20^th^ there were only 21 new cases and two new deaths. These numbers are so small that slight increases would have no noticeable effect on the ratio of cumulative deaths/cumulative cases in mid-April. In addition, deaths are unlikely to be underestimated because South Korea tested all suspected cases and attributed all deaths of persons who tested positive for the coronavirus to the virus, even if they had other underlying health conditions [AFP-JiJi, 2020].

The average death rate for COVID-19 in a country is very sensitive to the age distribution of infected individuals in a country because death rates are much higher for older patients. The South Korean death rate is only applicable for the U.S. if the age distributions of COVID-19 patients are similar in both countries.

The age distribution of all cases is known in South Korea, but it is not known in the U.S. because such a small fraction of the COVID-19 cases have been tested. The distributions of the known cases in the U.S. in mid-April includes 40% in the over-60 category compared to 25% in South Korea [Breton, 2020]. The higher reported incidence in the U.S. is due, at least in part, to the failure to test individuals who were asymptomatic or had mild symptoms, a category where younger individuals are more prevalent. Taking this characteristic into account, the age distributions of infected individuals do not seem very different in the two countries, so I conclude the South Korean death rate of 2.3% is applicable for the actual death rate in the U.S. when all cases of COVID-19 are included.

## III. U.S. Data on the Share of Positive Tests and the Associated Death Rates

Several groups track the numbers of U.S. tests, confirmed cases, and deaths from COVID-19 by state and publish their results several times a day. These data can be used to calculate the shares of individuals that test positive and the death rates associated with the confirmed cases.

The reported cases of COVID-19 in the U.S. in April 2020 are a subset of the actual number of cases because the U.S. did not have sufficient testing capacity to test all of the individuals with symptoms of the disease. Due to the limited tests available, the CDC issued guidelines to the states to prioritize testing [Connor, 2020]:

- Priority one: Hospitalized patients and symptomatic healthcare workers
- Priority two: High-risk patients with coronavirus symptoms
- Priority three: Symptomatic individuals in the community, if resources allow

None of these priorities included asymptomatic individuals.

The likelihood that tests would be positive was highest for individuals in Priority one and lower in Priorities two and three. As a result, as the number of tests increases, testing is extended to groups less likely to test positive, and the positive share of cumulative tests declines. States (or cities) overwhelmed with coronavirus cases tested only a small fraction of the individuals they thought were infected and obtained a very high share of positive tests.

Figure 1 shows the share of cumulative positive tests by state for the lower-48 states and South Korea on April 7^th^, using U.S. data published in *Politico* [Jin, 2020]. These data show that about half the states had shares of positive tests under 10%, with the lowest share at 3%. All of the states had a higher share of cumulative positive tests than South Korea (2%). Five states had shares of cumulative positive tests over 20%, and three states had shares over 30%. These three states, New Jersey, New York, and Michigan, reported tremendous limitations on their capacity to test individuals with COVID-19 symptoms.

**Figure 1.**
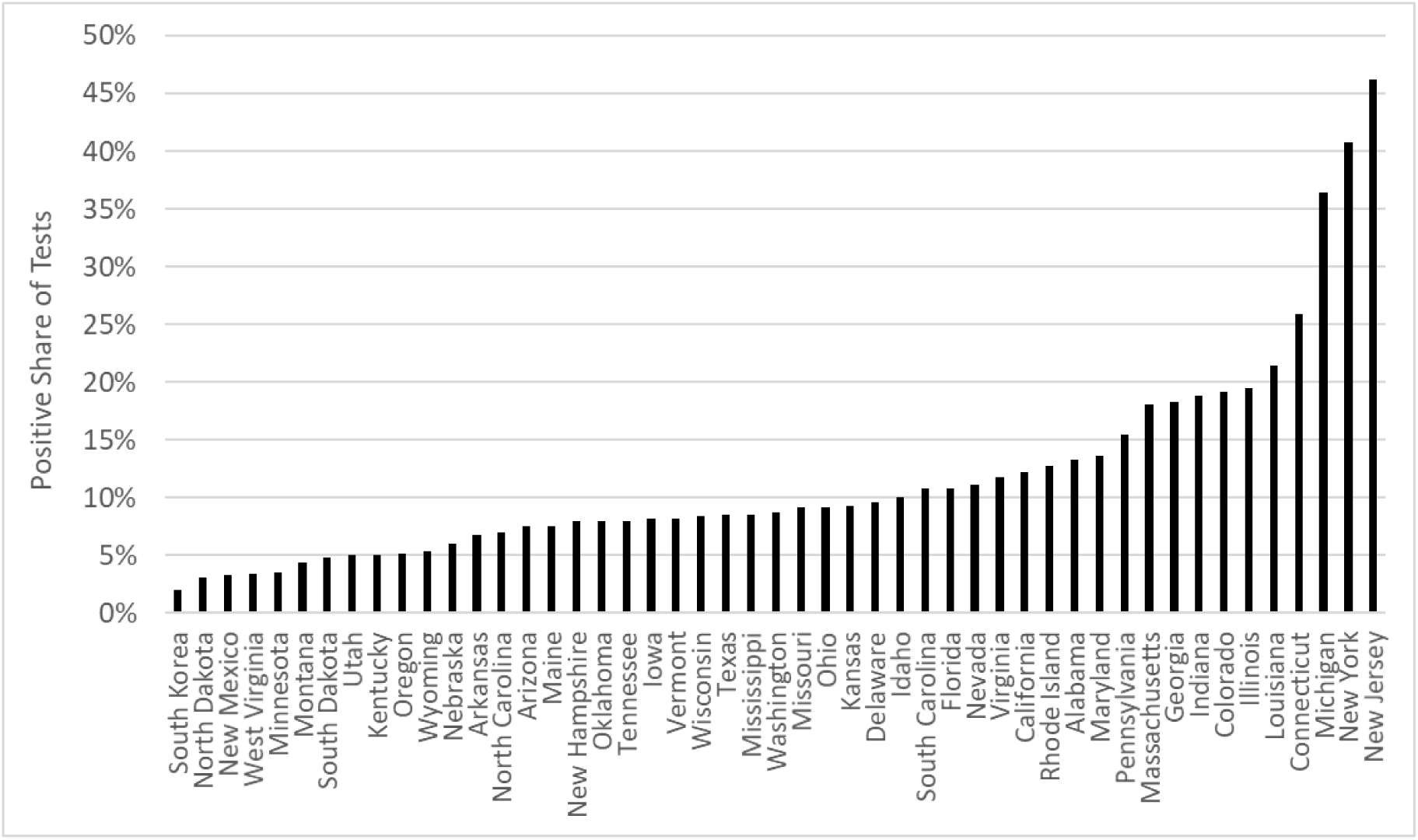
Positive Share of Cumulative Coronavirus Tests on April 7, 2020.

Maag [2020] reports that in late March 2020 very sick New Jersey residents spent many nights in their cars waiting in line at testing stations, after being turned away in emergency rooms. In late March New York City restricted testing to hospitalized patients to prevent the many infected individuals with less serious symptoms from leaving home [Cuzey, 2020]. Michigan restricted testing to those with the most serious symptoms until testing was extended to those with mild symptoms in mid-April [Clarke, 2020].

Until mid-March even states with lower shares of cumulative positive tests, such as California and Rhode Island, reported that they could not test everyone who needed a test [Becker, 2020 and Mooney, 2020]. Even in mid-April, after testing capacity in the U.S. improved relative to the number of suspected COVID-19 cases, tests in most locations were still rationed to the higher priority applications [Connor, 2020].

Death rates (deaths/infected population) for COVID-19 calculated from the confirmed cases for most states are much higher than the 2.3% rate in South Korea, even though there is no reason to expect that a higher share of Americans is likely to die from the virus. The most likely explanation for the higher death rates in the U.S. is that a large share of infected individuals are not included in the data.

If this is the case, then we should also expect to see a strong positive correlation between the death rates and the share of cumulative positive tests across states. A higher share of individuals testing positive is almost certain to indicate that the COVID-19 cases more difficult to identify are missing.

Figure 2 shows a plot of the coronavirus death rate from cumulative reported cases and the share of cumulative positive tests for reported cases in the lower-48 states on April 7, 2020. I use Jin’s [2020] data on cumulative tests and positive test results by state on April 7^th^ and the New York Times’ [2020] data on cumulative deaths by state on April 21^st^ to calculate the share of positive tests and the death rates for the lower-48 states.

**Figure 2.**
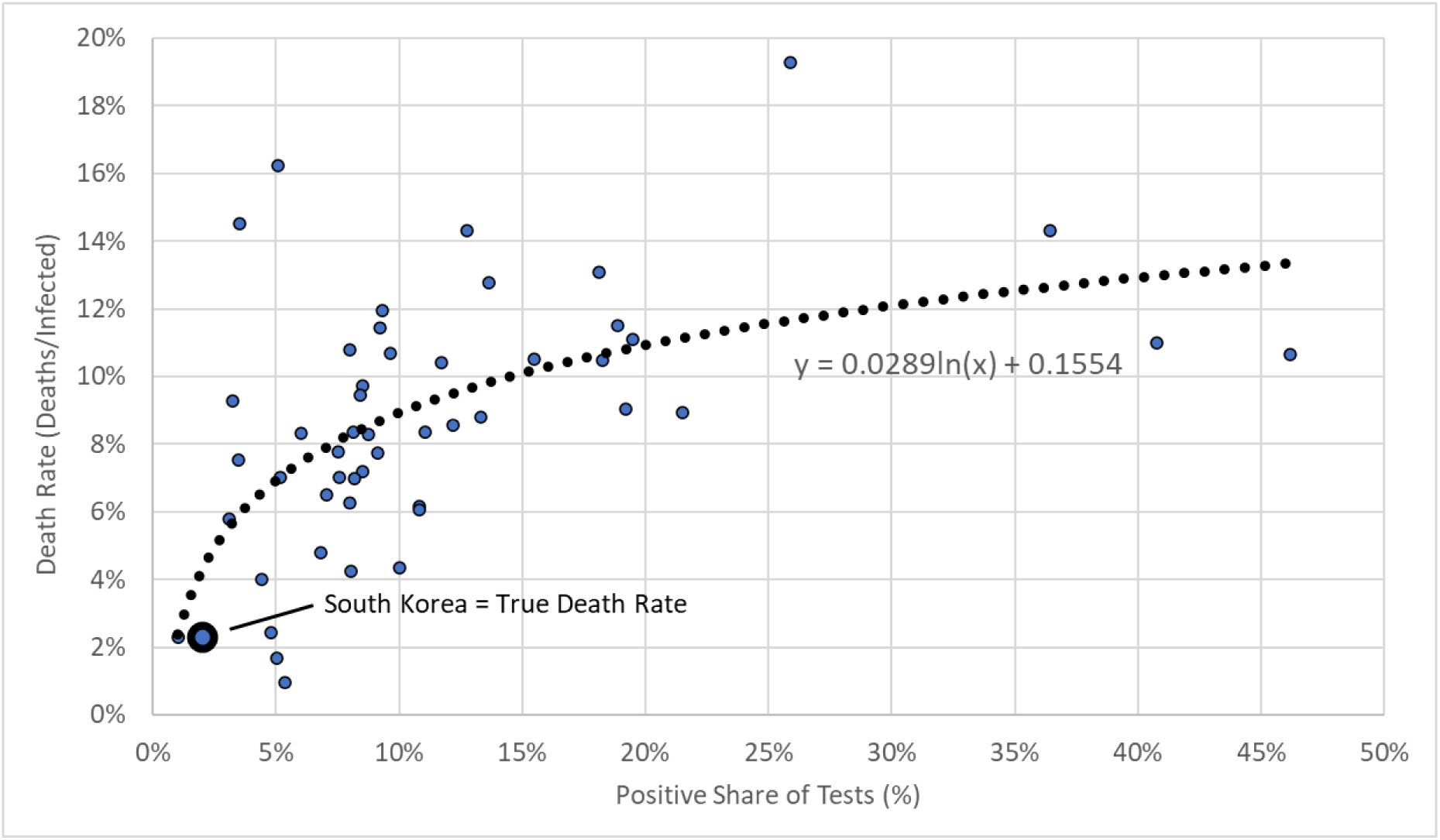
Apparent U.S. Death Rates vs. Positive Shares of Tests on April 7th.

The reported lag for test results from LabCorp and Quest Diagnostics on April 8^th^ was 3-4 days [Strickler and Kaplan, 2020]. With a total lag of 19 days between the onset of symptoms and death, I assume that the average time lag between the onset of symptoms and test results on April 7^th^ was 5 days. This leaves a 14-day average lag between reports of cases of coronavirus and the deaths associated with these cases.

The data for the death rate and the positive share of tests in the figure show a definite positive logarithmic relationship (R^2^ = 0.30). As the positive share of cumulative tests rises across states, the associated death rate rises, but at a decreasing rate. The share of cumulative positive tests in the trendline associated with the assumed 2.3% true death rate is 1%, which in the absence of tracking, is consistent with the 2% share of positive tests at that death rate in South Korea.

The estimated relationship of the death rate (deaths/infected population) to the positive share of cumulative tests is:

1. Death rate = 0.0289 ln(positive share of tests) + 0.155

The data used to estimate this relationship are provided in Breton [2020].

This relationship can be used to estimate the ratio of the actual infected population to the identified cases when the death rate equals 2.3% as a function of the positive share of cumulative tests in each state:

2) TotaI Cases/Identified Cases = 1.257 ln(positive share of tests) + 6.757

Equation (2) can be used to estimate the total number of actual COVID-19 cases in each state or in a subregion of a state. The sum of all the actual cases in the states can be used to estimate the infected share of the U.S. population.

Equation (2) is based on the assumption that the reported deaths due to COVID-19 in the U.S. are as complete as the reported deaths in South Korea. As discussed earlier, South Korea tracked and tested all individuals likely to have COVOD-19 and attributed any deaths of patients who tested positive for COVID-19 to the virus. This is not the case in the U.S. because some deaths from the virus involved patients who died at home or in nursing homes without ever being tested for the virus. These deaths were attributed to pneumonia, to some underlying condition, or to some fatal physiological response to the virus, such as a stroke.

Coroners are now reviewing deaths that occurred over the last few months and increasing the deaths attributed to COVID-19, but these deaths are not included in the *New York Times* [2020] data. This means that the death rate in Equation (1) and the ratio of total cases/identified cases in Equation (2) are underestimated.

Wu, McCann, Katz, and Peltier [2020] have compared the surge in mortality in New York City from March 11, 2020 to April 25, 2020 (relative to the average for the same period in 2017-2019) to the number of COVID-19 deaths in the *New York Times* data. The sudden increase in deaths in 2020 is 25% higher (4200/16673) than the reported COVID-19 deaths, which could be unreported COVID-19 deaths. They show similar trends in numerous European countries.

Given this information, the estimates from Equations (1) and (2) should be considered a lower bound on U.S. COVID-19 death rates and on the ratio of total/reported cases of COVID-19. A more reasonable estimate of these two variables appears to be a range from the values estimated by the Equations to values that are 25% higher.

The validity of this estimated range for total COVID-19 cases in each state can be evaluated by examining the plausibility of the estimated unidentified shares of cases in the states with the lowest and highest shares of positive tests in April, given the reported availability of tests in those states in late March and early April.

Applying Equation (2) and a value 25% higher in Montana, the state with the lowest share of positive tests (3.7%) on April 25th, indicates that the state identified 31-38% of its COVID-19 cases. Since 38% of all cases are estimated to be asymptomatic, if the state did not identify any of these cases, then it identified 50-61% of the symptomatic cases. Since most cases are mild or moderate, with symptoms not obviously due to Covid-19, this estimate seems reasonable, even given the relatively high level of testing in the state.

Applying Equation (2) and a value 25% higher in New Jersey, the state with the highest share of positive tests (49.6%) on April 25th, indicates that the state identified only 14-17% of its COVID-19 cases. Since 38% of the cases are estimated to be asymptomatic, if the state did not identify any of these cases, then it identified 23-27% of the symptomatic cases. Given the tremendous difficulty encountered by individuals seeking tests in New Jersey in late March, and the likelihood that a portion of the severe cases ending in death were not identified, this estimate also seems reasonable.

## IV. Estimated Total and Unidentified Coronavirus Cases

Table 2 presents the results for an application of the model and values 25% higher in each state, using Jin’s [2020] data for the share of cumulative positive tests on April 25^th^. The state results are used to calculate results for the U.S. as a whole, which due to the lag in testing and obtaining test results, measure the levels of infection about 10 days earlier. For each state the table shows the minimum ratio of total cases/reported cases, the minimum share of unidentified cases, and the range of infected shares of the population in mid-April.

**Table 2.** Unidentified COVID-19 Cases and Share of U.S. Population Infected in Mid-April 2020.

| States | Positive Tests | Tests | Positive Share | Min Tot /Ident | Min Tot Cases | Min Not Identified | Population | Infected Min | Infected Max |
| --- | --- | --- | --- | --- | --- | --- | --- | --- | --- |
| Alabama | 5,832 | 52,695 | 11.1% | 3.99 | 23,271 | 74.9% | 4,858,979 | 0.5% | 0.6% |
| Arizona | 6,045 | 60,714 | 10.0% | 3.86 | 23,317 | 74.1% | 6,828,065 | 0.3% | 0.4% |
| Arkansas | 2,741 | 35,578 | 7.7% | 3.53 | 9,689 | 71.7% | 2,978,204 | 0.3% | 0.4% |
| California | 39,254 | 494,173 | 7.9% | 3.57 | 140,264 | 72.0% | 39,144,818 | 0.4% | 0.4% |
| Colorado | 11,262 | 52,324 | 21.5% | 4.83 | 54,353 | 79.3% | 5,456,574 | 1.0% | 1.2% |
| Connecticut | 23,921 | 74,038 | 32.3% | 5.34 | 127,662 | 81.3% | 3,590,886 | 3.6% | 4.4% |
| Delaware | 3,442 | 17,379 | 19.8% | 4.72 | 16,252 | 78.8% | 945,934 | 1.7% | 2.1% |
| Florida | 30,174 | 316,959 | 9.5% | 3.80 | 114,685 | 73.7% | 20,271,272 | 0.6% | 0.7% |
| Georgia | 22,147 | 107,176 | 20.7% | 4.77 | 105,,752 | 79.1% | 10,214,860 | 1.0% | 1.3% |
| Idaho | 1,836 | 19,091 | 9.6% | 3.81 | 7,002 | 73.8% | 1,654,930 | 0.4% | 0.5% |
| Illinois | 39,658 | 189,632 | 20.9% | 4.79 | 189,964 | 79.1% | 12,859,995 | 1.5% | 1.8% |
| Indiana | 13,680 | 75,553 | 18.1% | 4.61 | 63,050 | 78.3% | 6,619,680 | 1.0% | 1.2% |
| Iowa | 4,445 | 31,973 | 13.9% | 4.28 | 19,010 | 76.6% | 3,123,899 | 0.6% | 0.8% |
| Kansas | 2,777 | 23,588 | 11.8% | 4.07 | 11,296 | 75.4% | 2,911,641 | 0.4% | 0.5% |
| Kentucky | 3,481 | 42,844 | 8.1% | 3.60 | 12,537 | 72.2% | 4,425,092 | 0.3% | 0.4% |
| Louisiana | 26,140 | 143,716 | 18.2% | 4.61 | 120,626 | 78.3% | 4,670,724 | 2.6% | 3.2% |
| Maine | 965 | 17,749 | 5.4% | 3.10 | 2,988 | 67.7% | 1,329,328 | 0.2% | 0.3% |
| Maryland | 16,616 | 84,716 | 19.6% | 4.71 | 78,252 | 78.8% | 6,006,401 | 1.3% | 1.6% |
| Massachusetts | 46,023 | 195,076 | 23.6% | 4.94 | 227,426 | 79.8% | 6,794,422 | 3.3% | 4.2% |
| Michigan | 36,641 | 136,296 | 26.9% | 5.11 | 187,079 | 80.4% | 9,922,576 | 1.9% | 2.4% |
| Minnesota | 3,185 | 53,787 | 5.9% | 3.20 | 10,205 | 68.8% | 5,489,594 | 0.2% | 0.2% |
| Mississippi | 5,434 | 55,670 | 9.8% | 3.83 | 20,824 | 73.9% | 2,992,333 | 0.7% | 0.9% |
| Missouri | 6,625 | 65,207 | 10.2% | 3.88 | 25,722 | 74.2% | 6,083,672 | 0.4% | 0.5% |
| Montana | 444 | 12,127 | 3.7% | 2.60 | 1,154 | 61.5% | 1,032,949 | 0.1% | 0.1% |
| Nebraska | 2,124 | 18,612 | 11.4% | 4.03 | 8,557 | 75.2% | 1,896,190 | 0.5% | 0.6% |
| Nevada | 4,398 | 36,192 | 12.2% | 4.11 | 18,065 | 75.7% | 2,890,845 | 0.6% | 0.8% |
| New Hampshire | 1,670 | 16,809 | 9.9% | 3.85 | 6,437 | 74.1% | 1,330,608 | 0.5% | 0.6% |
| New Jersey | 102,196 | 205,962 | 49.6% | 5.88 | 600,513 | 83.0% | 8,958,013 | 6.7% | 8.4% |
| New Mexico | 2,379 | 46,563 | 5.1% | 3.02 | 7,181 | 66.9% | 2,085,109 | 0.3% | 0.4% |
| New York | 271,590 | 730,656 | 37.2% | 5.51 | 1,497,279 | 81.9% | 19,795,791 | 7.6% | 9.5% |
| North Carolina | 8,052 | 100,584 | 8.0% | 3.58 | 28,850 | 72.1% | 10,042,802 | 0.3% | 0.4% |
| North Dakota | 748 | 17,449 | 4.3% | 2.80 | 2,093 | 64.3% | 756,927 | 0.3% | 0.3% |
| Ohio | 14,581 | 107,109 | 13.6% | 4.25 | 61,975 | 76.5% | 11,613,423 | 0.5% | 0.7% |
| Oklahoma | 3,121 | 46,140 | 6.8% | 3.37 | 10,522 | 70.3% | 3,911,338 | 0.3% | 0.3% |
| Oregon | 2,177 | 45,492 | 4.8% | 2.94 | 6,392 | 65.9% | 4,028,977 | 0.2% | 0.2% |
| Pennsylvania | 38,652 | 186,143 | 20.8% | 4.78 | 184,799 | 79.1% | 12,802,503 | 1.4% | 1.8% |
| Rhode Island | 6,699 | 47,257 | 14.2% | 4.30 | 28,814 | 76.8% | 1,056,298 | 2.7% | 3.4% |
| South Carolina | 4,917 | 44,463 | 11.1% | 3.99 | 19,615 | 74.9% | 4,896,146 | 0.4% | 0.5% |
| South Dakota | 2,040 | 14,824 | 13.8% | 4.26 | 8,699 | 76.5% | 858,469 | 1.0% | 1.3% |
| Tennessee | 8,726 | 131,328 | 6.6% | 3.35 | 29,221 | 70.1% | 6,600,299 | 0.4% | 0.6% |
| Texas | 22,806 | 242,547 | 9.4% | 3.79 | 86,326 | 73.6% | 27,469,114 | 0.3% | 0.4% |
| Utah | 3,782 | 84,697 | 4.5% | 2.85 | 10,776 | 64.9% | 2,995,919 | 0.4% | 0.4% |
| Vermont | 827 | 14,310 | 5.8% | 3.17 | 2,624 | 68.5% | 626,042 | 0.4% | 0.5% |
| Virginia | 11,169 | 69,015 | 16.2% | 4.47 | 49,901 | 77.6% | 8,382,993 | 0.6% | 0.7% |
| Washington | 12,753 | 153,376 | 8.3% | 3.63 | 46,302 | 72.5% | 7,170,351 | 0.6% | 0.8% |
| West Virginia | 988 | 29,811 | 3.3% | 2.47 | 2,445 | 59.6% | 1,844,128 | 0.1% | 0.2% |
| Wisconsin | 5,356 | 59,929 | 8.9% | 3.72 | 19,932 | 73.1% | 5,771,337 | 0.3% | 0.4% |
| Wyoming | 349 | 8,045 | 4.3% | 2.81 | 982 | 64.4% | 586,107 | 0.2% | 0.2% |
| **Total** | **884,868** | **4,815,374** | **18.4%** | **4.89** | **4,330,679** | **79.6%** | **318,576,557** | **1.4%** | **1.7%** |
| New York City | 158,258 | 373,736 | 42.3% | 5.68 | 898,404 | 82.4% | 8,398,748 | 10.7% | 13.4% |

What is striking is that most of the cases in every state are unidentified. Overall, only 16-20% of all cases have been confirmed through testing, leaving 80-84% unidentified. The minimum unidentified share ranges from 60% to 83% across states. The total number of cases in the U.S. was 4.3-5.4 million, yielding an infected share of the population of 1.4-1.7%.

The rate of infection varies dramatically across states, from about 0.1% in Montana and West Virginia to a range of 7.6 to 9.5% in New York. Applying the model to New York City using testing data from New York State [2020] for April 25^th^, indicates that the rate of infection in the City in mid-April was 10.7-13.4%.

## V. Discussion

The U.S. testing to identify cases of COVID-19 has been completely inadequate to determine the extent of infection in the population. In the absence of sufficient tests, this study uses the death rates and the shares of cumulative positive tests in the lower-48 states and the death rate in South Korea to estimate the size of the population infected with COVID-19 and the share in each state that has been identified.

The results indicate that 1.4-1.7% of the U.S. population was infected with the virus in mid-April. This share is nowhere near enough to create herd immunity, but it is far too many people to permit contact tracing in the states with the highest infection rates. South Korea managed to control the spread of the virus through a contact tracing process, but they only had 10,600 total cases.

The results indicate that no state has managed to identify even half of the COVID-19 cases, so it is difficult to control the spread. But it is also clear that not all states have been severely affected by the virus, with rates of infection in mid-April ranging from as little as 0.1% in rural states to a high of 7.6-9.5% in New York State.

The wide differences in rates of infection indicate that different strategies are appropriate to manage the virus in different states. Contact tracing would be feasible, at least in theory, in the states with the lower rates of infection, as South Korea managed to do it with a 0.2% infected share of its population.

## Data Availability

Data will be available in an SSRN paper with Abstract ID 3583941, as soon as this paper is approved for posting.

